# Study Protocol for the Health Outcomes in Pregnancy and Early Childhood (HOPE) Study: A Mother-Infant Study in American Samoa

**DOI:** 10.1101/2025.06.04.25329013

**Authors:** Lacey W. Heinsberg, Miracle Loia, Susie Tasele, Kima Faasalele-Savusa, Jenna C. Carlson, Scott Anesi, Katie Desobry, Efren Yuchongco, Brian Fortuno Guevara, Aigaeiva Sesaga, Alice Iloilo, Va’atausili Tofaeono, Kaylynn Bryan, Tina Tauasosi-Posiulai, Erin E. Kershaw, Yvette P. Conley, Daniel E. Weeks, Nicola L. Hawley, Bethel Muasau-Howard

**Author notes:** Equal contributions as first author (Lacey W. Heinsberg, Miracle Loia). Equal contributions as senior author (Nicola L. Hawley, Bethel Muasau-Howard). **Authors Email addresses** Lacey W. Heinsberg Miracle Loia Susie Tasele Kima Faasalele-Savusa Jenna C. Carlson Scott Anesi Katie Desobry Efren Yuchongco Bian Guevara Aigaeiva Sesaga Alice Iloilo Va’atausili Tofaeono Kaylynn Bryan Tina Tauasosi-Posiulai Erin E. Kershaw Yvette P. Conley Daniel E. Weeks Nicola L. Hawley Bethel Muasau-Howard. **Authors contributions:**. **Corresponding author**: Lacey W. Heinsberg, PhD, RN, Department of Health Promotion and Development, School of Nursing, University of Pittsburgh, 440 Victoria Building, 3500 Victoria Street, Pittsburgh, PA 15261, |.

## Abstract

**Introduction:** Pacific Islanders, including those in American Samoa, face a disproportionately high burden of gestational diabetes mellitus (GDM) and related sequalae of metabolic conditions. The *CREBRF* rs373863828 genetic variant, which is uniquely common among Pacific Islanders, has been paradoxically associated with higher body mass index (BMI) but lower risk of type 2 diabetes. While emerging evidence suggests this variant may influence both maternal metabolic outcomes and infant growth, studies in pregnancy and early life remain limited. The purpose of this paper is to describe the protocol for a study designed to address these gaps.

**Methods and analysis:** The Health Outcomes in Pregnancy and Early Childhood (HOPE) Study is an observational, longitudinal cohort study that will enroll up to 180 Samoan pregnant women and their infants (target n=150 dyads completing study protocols) in American Samoa, with follow-up through six months postpartum/postnatal. The study includes questionnaires, anthropometric measurements, and biospecimen collection. Genetic and epigenetic analyses will examine associations between maternal and infant *CREBRF* rs373863828 genotype, gestational diabetes status, infant body size, and cord blood DNA methylation.

**Ethics and dissemination:** The study is approved by the Institutional Review Boards at the University of Pittsburgh, Yale University, and the American Samoa Department of Health, as well as the Lyndon B. Johnson Tropical Medical Center (American Samoa) Research Oversight Committee. Findings will be disseminated through peer-reviewed publications, conference presentations, and community reports.

## 1.0 INTRODUCTION

Pacific Islander women in the United States (U.S.) face a disproportionately higher risk of gestational diabetes mellitus (GDM) compared to women of European ancestry (9.9–14.8% versus 2–6%, respectively) (1). In American Samoa, GDM statistics far exceed these numbers, affecting up to 40% of Samoan women (2). GDM not only impacts maternal health but also birth outcomes and long-term health of the offspring (3,4). For example, children born to mothers with GDM have an increased risk of metabolic irregularities that can promote the intergenerational transmission of obesity and chronic disease (5,6).

The genetic marker *CREBRF* rs373863828 is receiving increasing attention for its unique health effects in Pacific Islander populations (7–9) and its potential to improve our understanding of GDM and related child health outcomes. The variant’s minor allele (A) is uniquely common among Pacific Islanders (minor allele frequency ∼0.26) and is associated with *greater* body mass index (BMI) but paradoxically *lower* odds of type 2 diabetes among adults (8). These associations have been replicated across multiple Pacific Islander populations (9–13), and early evidence suggests the variant may also protect against GDM (14).

Although research on early life, a critical window for long-term metabolic programming, is still limited, evidence suggests that *CREBRF* may also influence infant growth and body composition. In a sample of young children from Samoa recruited at birth and followed through 2 years, the presence of the A allele (AA/AG genotypes) was associated with greater bone and lean mass in infants (15) and greater height in toddlers (16) compared to those without the A allele (GG genotype), which may begin to explain its longer-term metabolic health benefits (17–19). Additional findings indicate that the association with height persists into early childhood and that, by around age four, associations with more clinically visible obesity-related phenotypes, such as greater weight and abdominal circumference, also begin to emerge (20).

Despite these findings, most *CREBRF* research has focused either on maternal genetic contributions to GDM or offspring genetic influences on growth, without considering the complex interplay between maternal-offspring genetics. This is an important gap because pregnancy is a time of dynamic biological adaptation during which the maternal metabolic system shifts to prioritize nutrient delivery for fetal development (21). This shift is thought to be shaped by the mother’s metabolic status and genetic makeup alongside hormonal factors produced by the placenta, which generally shares the same genetic profile as the fetus (22). This suggests that GDM may not emerge in isolation, but instead from suboptimal maternal-fetal resource allocation influenced by both maternal and fetal genetics (22,23). Fetal growth is similarly shaped by this dual influence—indirectly by maternal genetics, which govern the intrauterine environment (24), and directly by fetal genetics, which affect insulin and hormone production both before and after birth (25). In the case of *CREBRF*, we hypothesize that maternal-fetal co-occurrence of the A allele synergistically provides greater protection against GDM compared to maternal genetics alone, leading to improved anthropometric outcomes in children via fetal programming. Supporting this broader framework, other studies have shown that maternal-offspring genotype combinations (e.g., at *IRS1* rs1801278 (26)) can modulate GDM risk and, in turn, infant outcomes.

Beyond genetic contributions, epigenetic mechanisms such as DNA methylation may also play a key role in shaping infant health. As a critical regulator of gene expression and a central mechanism in fetal metabolic programming, DNA methylation is influenced by both genetic and environmental factors during fetal development (27,28). Given its importance in human health, it is essential to understand how maternal–infant *CREBRF* genotype combinations affect DNA methylation, and how these genetic effects may interact with metabolic exposures, such as GDM, to shape the infant epigenome. Likewise, and more broadly, it is also important to understand how DNA methylation at birth can impact infant growth and development as this may help identify biological pathways through which maternal-fetal genetic interactions and prenatal exposures influence long-term metabolic health.

To address these gaps, we have designed the Health Outcomes in Pregnancy and Early Childhood (HOPE) study, which aims to understand the health and wellness of American Samoan women and their children, with a specific focus on GDM and infant growth. The primary objective is to examine how maternal-infant *CREBRF* rs373863828 genotypes jointly contribute to GDM status, infant body size, and genome-wide DNA methylation in cord blood at birth. Secondary objectives include integrating social, environmental, and behavioral data to understand how these contextual factors shape health outcomes, as well as exploring additional contributors to maternal and child health in this population, including genetic (e.g., *BTNL9* rs200884524, another Pacific Islander-specific variant linked to cardiometabolic health (29)), genomic (e.g., microbiome composition, which may indirectly influence infant growth and immune development (30)), and environmental (e.g., per and polyfluoroalkyl substance concentrations, which have been linked to metabolic disruption (31)).

## 2.0 METHODS

### 2.1 Study design and overview

This ongoing, longitudinal, observational study (recruitment started April 2025) aims to enroll up to n=180 Samoan pregnant women and their index offspring from American Samoa, with a target of n=150 mother-infant dyads completing study protocols. Eligible women will be enrolled during their third trimester of pregnancy, and each dyad will be followed through the infant’s first six months of life. Participants will complete five study visits: one during pregnancy, one shortly after birth, and follow-up visits at 2, 4, and 6 months postpartum. As part of the study, mothers will complete questionnaires about themselves, their families, and their infants; physical measurements and biospecimens will be collected from both mothers and infants; and additional health data will be abstracted from medical records to supplement study assessments. Further details on study procedures are provided below.

### 2.2 Ethics

The HOPE study has received Institutional Review Board (IRB) approval from both the University of Pittsburgh and Yale University, with the University of Pittsburgh serving as the IRB of record through a multi-site IRB agreement (STUDY24020055). Local and territorial approval has been granted by the American Samoa IRB. In addition, the study has been approved by the Lyndon B Johnson Tropical Medical Center (LBJ) Research Oversight Committee for all hospital-related activities.

### 2.3 Setting

American Samoa is an unincorporated territory of the U.S. located approximately halfway between Hawaii and New Zealand in the South Pacific Ocean (32). It consists of several islands, the largest and most populated being Tutuila, which is home to >90% of residents (32). Tutuila measures roughly 21 miles in length and 3 miles across at the widest points.

The American Samoan population consists predominantly of Samoan individuals. Given its mountainous terrain, American Samoa is quite urbanized with the main population center being Tafuna, home to ∼8,000 residents (33). According to the U.S. Census Bureau, the 2020 population of American Samoa was 49,710, a decline from 55,519 in 2010 (34), reflecting significant out migration, largely to the mainland U.S., and a decline in birth rates. American Samoa was recently classified as a high-income economy by the World Bank (transition from upper-middle economy the previous year due to revision in population estimates based on the 2020 census (35)), although the gross national income of $18,017 per capita in 2022 (36) is far lower than the $78,035 in the U.S. as a whole (37).

American Samoa’s healthcare system, designated as medically underserved by the U.S. Health Resources and Services Administration (HRSA) (38), includes one tertiary care facility, the Lyndon B Johnson Tropical Medical Center (LBJ), and four federally qualified community health centers (FQHCs) operated by the Department of Health. Residents are automatically enrolled in Medicaid through a waiver under the Social Security Act, bypassing individual eligibility assessments (39).

The health landscape in American Samoa has been shaped by complex historical dynamics, including foreign influence and multifaceted internal political, economic, and social systems. For example, as U.S. nationals without full political representation—and therefore no direct voice in federal decisions—American Samoans face structural barriers that affect healthcare access and broader resource allocation. Historical shifts in food systems, coupled with limited local infrastructure and geographic isolation, have further contributed to persistent health challenges.

Despite these challenges, the Samoan cultural values of *fa’aSamoa*—the Samoan way— emphasizes respect, reciprocity, and family and community ties (40), offering a powerful foundation for the future. This study, in partnership with the American Samoan community, incorporates Samoan frameworks such as *Teu le* va (maintaining respectful relationships (41)) and *Talanoa* (“talk story”, a dialogic approach rooted in storytelling (42,43)). By grounding the research in these values, we aim to ensure that knowledge sharing between researchers and the Samoan community occurs in a respectful and culturally safe environment, and that the resulting solutions to improve health outcomes for women and children are aligned with Samoan norms.

### 2.4 Participants

HOPE study participants will be recruited from prenatal care clinics and through flyers, social media advertising, and clinician referrals. Inclusion criteria for dyads include: (1) maternal age ≥18 years; (2) maternal report that the child has four Samoan grandparents (an empirically validated method of assessing ancestry in this setting (8), included due to the genetic focus of this study); (3) pregnancy gestation at or beyond 35 weeks (to limit heterogeneity related to extremely preterm birth); (4) singleton pregnancy (to avoid heterogeneity in growth patterns seen in multiples); (5) maternal plans to give birth at LBJ and reside in American Samoa for at least six months post-birth (to enable follow-up); (6) maternal completion of a standard-of-care 2-hour, 75g oral glucose tolerance test for GDM screening between 24 and 28 weeks’ gestation (to have a current best practice measure of GDM status); and (7) maternal intent to provide cord blood and saliva DNA specimens (to conserve limited study resources for the primary study goal, though participants are reminded at each visit that their involvement in each specific assessment is optional).

Exclusion criteria for dyads include: (a) pre-pregnancy maternal diabetes, defined by either self-report or a fasting glucose level of ≥126 mg/dL or HbA1c ≥6.5% during a standard prenatal visit before 12 weeks’ gestation; (b) maternal medical history of conditions that may interfere with maternal weight gain or fetal growth, such as prior bariatric surgery or congenital anomalies; or (c) insulin use as a first-line treatment for GDM (to reduce treatment-related heterogeneity).

For those eligible and interested, an orientation visit will be conducted by bilingual (English and Samoan) research assistants. Participant understanding will be assessed, and informed written consent obtained for enrollment of the pregnant woman and her unborn child. Participants receive incentives for each visit that they complete.

### 2.5 Study procedures

The HOPE study consists of five study visits: one before birth (Visit 1), one after birth (Visit 2), and follow-ups at 2, 4, and 6 months (Visits 3-5); the schedule of visit-specific research activities is outlined in Table 1. All data are collected by trained Samoan research team members who are bilingual in Samoan and English.

**Table 1.** Study timeline and research activities overview.

| **Table 1a. Maternal data collection** |  |  |  |  |  |  |
| --- | --- | --- | --- | --- | --- | --- |
|  | **Visit 1**<br>**(Before**<br>**birth)** | **Infant**<br>**birth** | **Visit 2**<br>**(After**<br>**birth)** | **Visit**<br>**3**<br>**(2mo)** | **Visit**<br>**4**<br>**(4mo)** | **Visit**<br>**5**<br>**(6mo)** |
| Saliva sampling | X |  |  |  |  | X |
| Capillary blood sampling | X |  |  |  |  | X |
| Physical measurements (height, weight, heart rate, blood pressure, and point-of-care HbA1c) | X |  |  | X | X | X |
| Body composition measurements |  |  |  | X | X | X |
| **Table 1b. Infant data collection** |  |  |  |  |  |  |
|  | **Visit 1**<br>**(Before**<br>**birth)** | **Infant**<br>**birth** | **Visit 2**<br>**(After**<br>**birth)** | **Visit**<br>**3**<br>**(2mo)** | **Visit**<br>**4**<br>**(4mo)** | **Visit**<br>**5**<br>**(6mo)** |
| Umbilical cord blood sampling |  | X |  |  |  |  |
| Saliva sampling |  |  | X |  |  | X |
| Capillary blood sampling |  |  | X | X |  | X |
| Stool sampling |  |  | X | X |  | X |
| Physical measurements (weight, length, head/abdominal circumference, skinfold thicknesses) |  |  | X | X | X | X |
| **Table 1c. Dyad and family data collection** |  |  |  |  |  |  |
|  | **Visit 1**<br>**(Before**<br>**birth)** | **Infant**<br>**birth** | **Visit 2**<br>**(After**<br>**birth)** | **Visit**<br>**3**<br>**(2mo)** | **Visit**<br>**4**<br>**(4mo)** | **Visit**<br>**5**<br>**(6mo)** |
| Water sampling (tap water, from the home) |  |  |  | X |  |  |
| Questionnaire administration | X |  | X | X | X | X |

Visit 1: Will occur between 35 weeks gestation and the onset of labor; Visit 2: Will occur within 0–7 days after birth; Visits 3-5: Will occur at approximately 2, 4, and 6 months of infant age, with a $\pm 4$ -week window around each target month. Note: Biospecimen collection not scheduled to occur at the 4 month visit to reduce participant burden.

#### 2.5.1 Physical (anthropometric and clinical) measurements

At Visits 1, 3, 4, and 5, research assistants will collect maternal height, weight, heart rate, blood pressure, and HbA1c measurements. Maternal height will be measured using a portable stadiometer (SECA, Hamburg, Germany), and weight will be recorded with a digital scale (Tanita Corporation of America, IL, USA) with participants wearing lightweight clothing. Anthropometric measurements will be taken in duplicate and averaged for analyses. Blood pressure and heart rate will be measured after a 10-minute seated rest period, with a 3-minute rest between readings, using an automated blood pressure monitor (Omron Healthcare). Three measurements will be taken and the last two will be averaged for analyses. A point-of-care device (PTS Diagnostics A1cNow+™ Systems) will be used to measure HbA1c via capillary blood collected from a finger stick after the finger is cleaned with an alcohol swab and allowed to dry. In the postpartum period only (Visits 3, 4, and 5), maternal body composition will be assessed via bioelectrical impedance analysis (Omron HBF-306C) to estimate fat mass and body fat percentage; this will be performed in only those participants without metal implants or pacemakers.

For infants, anthropometric measurements will be collected during Visits 2 through 5. Infant length and weight will be assessed with a length board and digital scale (SECA, Hamburg, Germany), with infants weighed in clean diapers after zeroing the scale for diaper weight. Head circumference will be measured at the widest possible point, and abdominal circumference will be measured above the belly button to avoid the umbilical cord stump in early assessments, using a standard tape measure (SECA, Hamburg, Germany). Measurements will be taken in duplicate and averaged for analyses. Age- and sex-standardized BMI z-scores will be calculated based on World Health Organization Child Growth Standards (44,45). Additionally, abdominal circumference-to-length ratio will be calculated as an indicator of abdominal visceral fat, determined by dividing abdominal circumference by length. Skinfold thicknesses at the tricep, bicep, subscapular, iliac crest, and thigh will be measured on the left side of the body using a Harpenden caliper (West Sussex, UK), and subcutaneous fat mass (mm²) will be estimated from the sum of all skinfold measurements.

#### 2.5.2 Biospecimen and environmental sample collection

##### Saliva

Saliva specimens will be collected from both mothers and infants using Oragene saliva collection kits for DNA and RNA stabilization (DNA Genotek). Mothers will provide saliva by spitting directly into the tube (DNA, OGR-600) or swabbing their mouth using a kit-supplied absorbent sponge (RNA, ORE-100). Infant saliva specimens will be collected only using a kit-supplied absorbent sponge (DNA, OC-175; RNA ORE-100). Once collected, the samples will be mixed with the stabilizing reagent within the tubes to ensure DNA and RNA integrity. Specimens will be securely stored at room temperature until they are shipped in batches to the University of Pittsburgh for DNA and RNA extraction and purification following standard protocols (DNA Genotek). Extracted DNA from maternal and infant saliva samples will be analyzed for *CREBRF* rs373863828 and *BTNL9* rs200884524 using established TaqMan® assays (Applied Biosystems).

##### Umbilical cord blood

Umbilical cord blood specimens will be collected immediately following delivery by trained staff at LBJ Hospital. After clamping and cleaning the umbilical cord, approximately 6 mL of blood will be drawn from the umbilical vein using an 18-gauge needle and sterile syringe. The specimen will then be aseptically transferred into PAXgene Blood DNA tube (Fisher Scientific, Catalog B761165, 2.5 mL) and a PAXgene Blood RNA tube (Fisher Scientific, Catalog 2302101, 2.5 mL). Samples will be frozen at -20°C for 24 hours and then transferred to -80°C storage for long-term preservation. Specimens will be shipped in batches on dry ice to the University of Pittsburgh for DNA and RNA extraction, analysis, and storage. Extracted DNA from cord blood will undergo epigenome-wide DNA methylation analysis using the EPIC v2.0 chip (Illumina).

##### Secondary samples

Several secondary samples will be collected and banked for future research. Capillary blood samples will be obtained from mothers (finger stick) and infants (heel stick) using sterile lancets and Mitra devices (Trajan Scientific Americas Inc.), with a total volume of up to 120 microliters per sample. The filled devices will be stored at -80°C and batch-shipped to the University of Pittsburgh for future per- and polyfluoroalkyl substance profiling. Infant fecal specimens will be collected using OMNIgene-gut kits (OMR-200, DNA Genotek) following manufacturer protocols, with stool obtained from used diapers before being stored at -80°C for future microbiome profiling. Finally, home water samples will be collected in pre-cleaned polypropylene bottles and stored at -20°C for future per- and polyfluoroalkyl substance profiling. Samples will be de-identified to protect participant confidentiality.

#### 2.5.3 Study questionnaires

Participants will complete a series of questionnaires (available in both English and Samoan, side-by-side) that gather information across several key domains including demographic, health, social, behavioral, and environmental factors. Demographic and household questions include topics such as education, relationship status, income, and other family and living characteristics. Health-related questions focus on pregnancy history, pre-pregnancy health, and pregnancy-related conditions, addressing both physical and psychosocial aspects. Behavioral and lifestyle questionnaires assess social support, tobacco and alcohol use, physical activity, nutrition, and sleep health. Environmental questions explore household and community context. Infant-focused sections capture information on feeding practices, sleep patterns, and developmental milestones. Participants will also be invited to share feedback on their study experience and indicate whether they are interested in receiving their genetic results. Additional details about the questionnaires are provided in Table 2. All questionnaire data will be collected using REDCap, a secure, web-based platform hosted on password-protected institutional servers (46,47). Data will be de-identified to protect participant confidentiality.

**Table 2.**
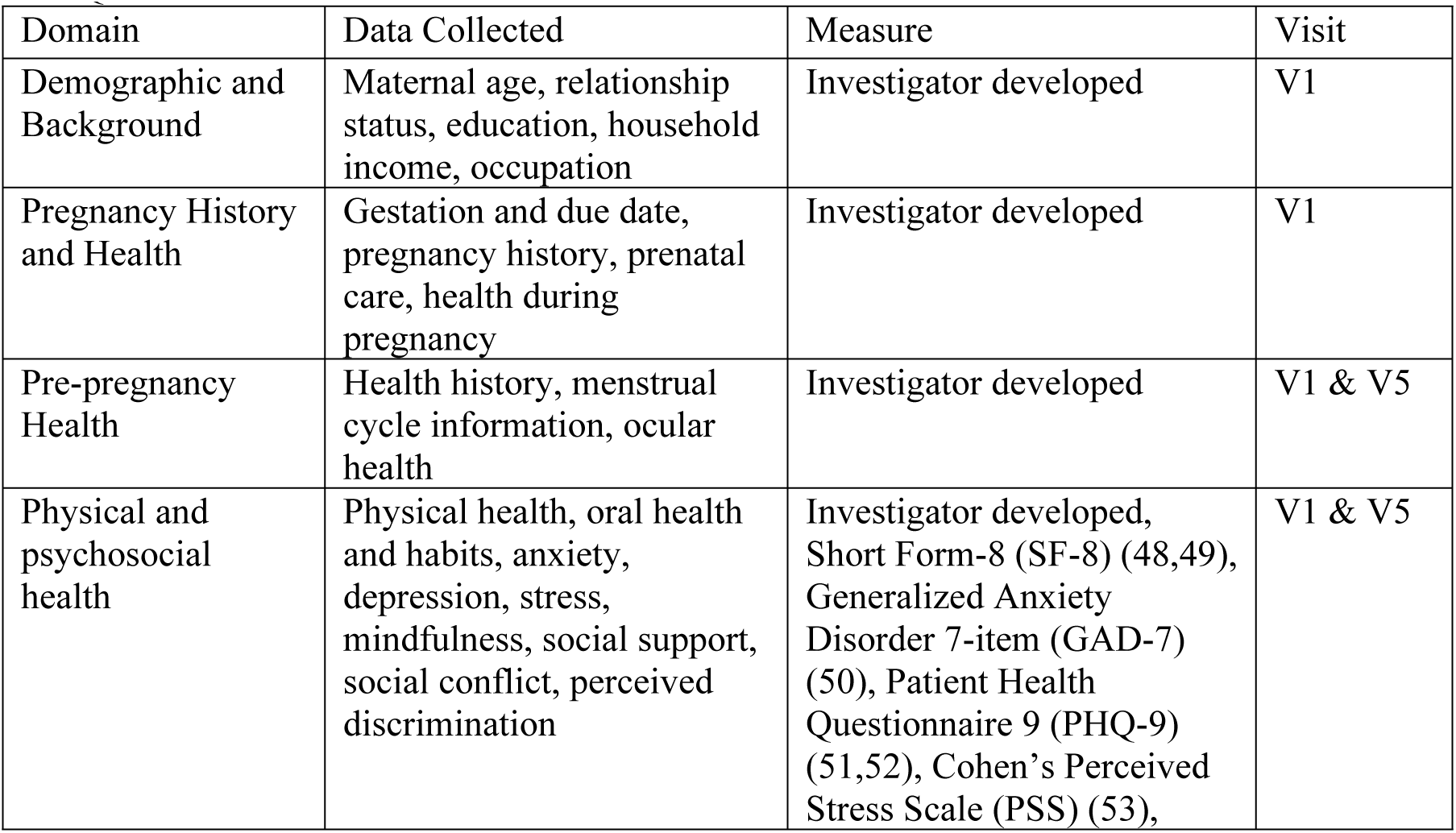
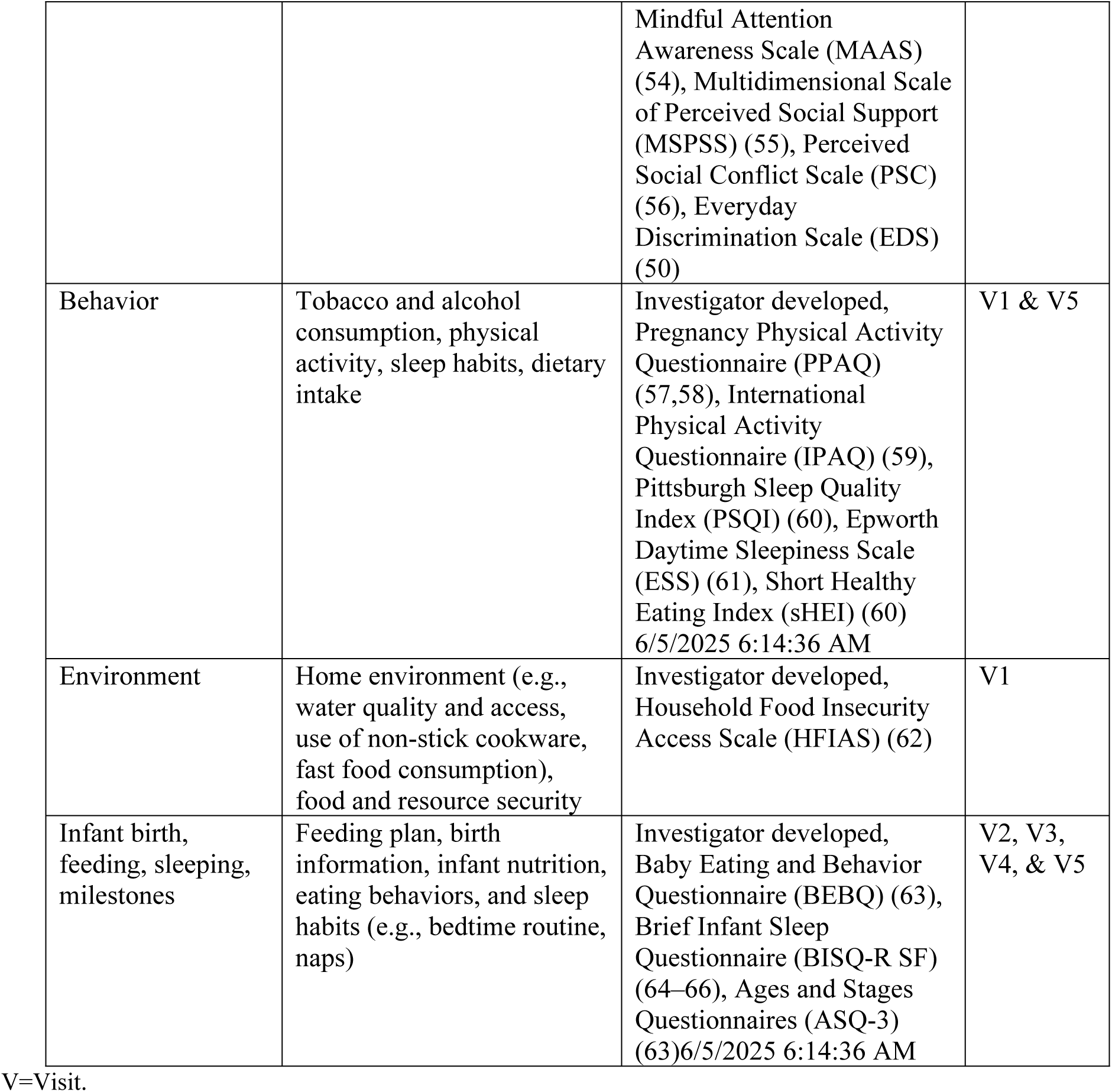
Questionnaires.

| Domain | Data Collected | Measure | Visit |
| --- | --- | --- | --- |
| Demographic and Background | Maternal age, relationship status, education, household income, occupation | Investigator developed | V1 |
| Pregnancy History and Health | Gestation and due date, pregnancy history, prenatal care, health during pregnancy | Investigator developed | V1 |
| Pre-pregnancy Health | Health history, menstrual cycle information, ocular health | Investigator developed | V1 & V5 |
| Physical and psychosocial health | Physical health, oral health and habits, anxiety, depression, stress, mindfulness, social support, social conflict, perceived discrimination | Investigator developed, Short Form-8 (SF-8) (48,49), Generalized Anxiety Disorder 7-item (GAD-7) (50), Patient Health Questionnaire 9 (PHQ-9) (51,52), Cohen's Perceived Stress Scale (PSS) (53), | V1 & V5 |
|  |  | Mindful Attention Awareness Scale (MAAS) (54), Multidimensional Scale of Perceived Social Support (MSPSS) (55), Perceived Social Conflict Scale (PSC) (56), Everyday Discrimination Scale (EDS) (50) |  |
| Behavior | Tobacco and alcohol consumption, physical activity, sleep habits, dietary intake | Investigator developed, Pregnancy Physical Activity Questionnaire (PPAQ) (57,58), International Physical Activity Questionnaire (IPAQ) (59), Pittsburgh Sleep Quality Index (PSQI) (60), Epworth Daytime Sleepiness Scale (ESS) (61), Short Healthy Eating Index (sHEI) (60)<br>6/5/2025 6:14:36 AM | V1 & V5 |
| Environment | Home environment (e.g., water quality and access, use of non-stick cookware, fast food consumption), food and resource security | Investigator developed, Household Food Insecurity Access Scale (HFIAS) (62) | V1 |
| Infant birth, feeding, sleeping, milestones | Feeding plan, birth information, infant nutrition, eating behaviors, and sleep habits (e.g., bedtime routine, naps) | Investigator developed, Baby Eating and Behavior Questionnaire (BEBQ) (63), Brief Infant Sleep Questionnaire (BISQ-R SF) (64–66), Ages and Stages Questionnaires (ASQ-3) (63)<br>6/5/2025 6:14:36 AM | V2, V3, V4, & V5 |

V=Visit.

#### 2.5.4 Medical record review and extraction

Clinical data will be extracted from participants’ medical records to supplement questionnaire responses and provide additional context. Extracted information will include GDM status, laboratory results (e.g., lipid levels), medication use, anthropometric measurements, birth-related details, and infant growth. Only personnel who are authorized, trained in privacy and data security (including the Health Insurance Portability and Accountability Act), and approved by LBJ Hospital will be permitted to access and extract this information.

#### 2.5.5 Participant feedback and referrals to clinical services

Study-related measurements will not be added to the participants’ medical records. However, participants may choose to receive a personalized “results booklet,” which will be maintained and updated throughout the study for those who opt in. This booklet will include maternal physical measurements (e.g., weight, height, heart rate, blood pressure, and HbA1c) and infant measurements (e.g., weight, length, head circumference, and abdominal circumference) and growth curves (using WHO growth charts). Before providing this information, research assistants will emphasize that all measurements are being collected for research purposes only and do not replace clinical care.

For unexpected health findings, participants will be referred to local clinicians at LBJ Hospital. Referral criteria include HbA1c levels ≥6.5%, blood pressure >140/90 mmHg (local referral threshold), and severe depression symptoms (Patient Health Questionnaire-9 (PHQ-9) score >20 or thoughts of self-harm). In cases of severe hypertension (≥160/110 mmHg) during pregnancy, an urgent preeclampsia evaluation referral will be provided.

Upon study completion, participants may request their own and/or their infant’s *CREBRF* rs373863828 and/or *BTNL9* rs200884524 genotype results. If requested, study staff will reinforce that these results are for research purposes only, may not be fully accurate, and cannot replace certified clinical testing. Participants will also be informed that the genetic data are still being studied and may not be directly useful at this time. Recommendations for maintaining a healthy lifestyle remain the same regardless of genotype.

### 2.6 Data management and statistical analysis

#### 2.6.1 Data management, cleaning, and quality control

All study data will undergo rigorous quality control procedures to ensure accuracy, consistency, and reliability. During data collection, quality assurance will include weekly to monthly reports, data capture review, and routine checks of data entry and protocol adherence. Automated validation rules and branching logic will be used within REDCap to help minimize entry errors in real-time. All modifications to study records will be documented through REDCap’s audit trail functionality. Data management includes secure storage on encrypted, password-protected servers, version-controlled datasets, and restricted access to identifiable information.

During the analysis phase, data quality diagnostics will be conducted to identify outliers, assess variable distributions, evaluate associations among variables, and examine missing data patterns. If needed, multiple imputation and sensitivity analyses will be employed to assess the impact of missingness.

For omics data, potential biases will be addressed through standardized laboratory controls, including the use of technical replicates, batch-specific standards, and internal efficiency measures. Preprocessing will include batch correction and other normalization techniques to ensure comparability and reduce technical variability.

#### 2.6.2 Data analyses

Primary statistical analyses will examine associations between genetic variation and health outcomes in mothers and infants, as well as characterize the infant cord blood epigenome (DNA methylation). This includes evaluating the impact of the *CREBRF* rs37386382 genotype on maternal outcomes (e.g., GDM status, blood pressure, HbA1c, and body size) and infant outcomes (e.g., birth size and body composition). Linear regression models will be used for continuous outcomes (e.g., anthropometric measures), while binary logistic regression will be used for binary outcomes (e.g., GDM status), adjusting for relevant covariates such as maternal age, BMI, and gestational age at birth. While the primary focus is on maternal-infant genotype combinations, maternal and infant genotypes will also be analyzed separately and in joint models to evaluate their individual, additive, and interactive effects on metabolic outcomes.

For DNA methylation analyses, epigenome-wide association studies (EWAS) will be conducted to assess associations between DNAm with genotype/genotype combinations (primary) and participant characteristics (secondary). M values (logit-transformed beta values) will be used in linear regression models to identify site-specific associations, adjusting for appropriate covariates. Analyses will be performed both with and without adjustment for cell type heterogeneity (67) inferred using deconvolution methods (68,69). Multiple testing correction will be applied using a methylome-wide significance threshold (p<9×10⁻⁸). Differentially methylated regions (DMRs) will be identified (70), and gene set enrichment analysis (GSEA) will be used to explore biological pathways associated with observed DNA methylation variation. Additional exploratory analyses are planned to support secondary study goals and future sample collections.

#### 2.6.3 Power

The study sample of up to 180 maternal-infant dyads (accounting for anticipated attrition to achieve a final analytic sample of approximately n=150 dyads) provides adequate power for the planned analyses. With n=150 dyads, we have ≥80% power to detect a moderate odds ratio (OR) of 1.75 for the association between maternal-offspring genotype combination and GDM, and a small to moderate effect size (Cohen’s d = 0.28) for associations with continuous infant anthropometric measures. Power calculations for DNA methylation analyses indicate sufficient sensitivity to detect moderate differences in DNA methylation levels at individual CpG sites at genome-wide significance thresholds.

### 2.7 Study status and timeline

Participant recruitment for HOPE began in April 2025. Recruitment is expected to be completed within 12–18 months, with follow-up visits anticipated to conclude within 18–24 months.

## 3.0 DISCUSSION

This study will support a hypothesis-driven examination of how maternal and infant genetics— particularly the *CREBRF* rs373863828 variant—relate to maternal and infant health in American Samoa. This work is significant because, unlike most genetic variants which have small effect sizes and are often overshadowed by social and behavioral factors, the *CREBRF* variant has demonstrated large and consistent effects (7,8,12,14,71), suggesting potential as a future screening or intervention target. As such, this study may help identify biological factors that contribute to health outcomes among the Pacific Islander population group which may inform population-specific strategies to improve outcomes. Additionally, the study is designed to contribute novel epigenetic and other omic data, offering valuable insight into potential mechanisms linking maternal and infant factors to health outcomes. Importantly, while this variant is enriched in Pacific Islander populations, its involvement in key metabolic pathways suggests broader relevance for understanding metabolic health in diverse populations.

Despite its significance, the study has some limitations. For example, recruitment is limited to the third trimester of pregnancy, restricting our ability to examine early pregnancy or pre-conception factors. Similarly, the study also does not include paternal data, preventing evaluation of extended family health behaviors or factors that may influence outcomes. In addition, while the use of non-invasive biospecimens was designed to reduce participant burden and distress and increase compliance, it may constrain the types of biomarkers that can be assessed. Finally, the study follows infants only through six months of age, limiting the ability to assess longer-term developmental trajectories.

Despite these constraints, several key strengths enhance the rigor and potential impact of this study. The focus of this work on a historically excluded population group with a high rate of GDM will ensure that findings contribute to reducing health disparities and improving the representation of Pacific Islander communities in genetic and environmental health research. In addition, the longitudinal design supports a comprehensive assessment of factors that shape early infant growth. Finally, the multimodal data collection approach including questionnaires, biospecimens, and genetic and epigenetic analyses enhances the study’s ability to investigate multiple biological pathways influencing maternal and infant health.

Therefore, this study represents a critical first step toward a biologically and culturally grounded understanding of maternal/child health outcomes in American Samoa. It lays the groundwork for future studies that may include earlier pregnancy recruitment, more detailed assessments of environmental and behavioral exposures, and extended follow-up into childhood. Taken together, this line of research may help guide the development of targeted, community-informed interventions to promote maternal and child health in American Samoa and other underrepresented populations globally.

## Data Availability

This is a study protocol paper--data collection for this study has just begun. However, participants in this study will be optionally consented for broad data sharing in federal registries. For those who consent, data will be available in dbGaP under accession number phs003874.v1.p1.

## DECLARATIONS

### Availability of data and materials

Participants in this study will be optionally consented for broad data sharing in federal registries. For those who consent, data will be available in dbGaP under accession number phs003874.v1.p1.

### Funding

This work is supported by the National Institutes of Health under award number K99HD107030, R00HD107030, faculty startup funds at the University of Pittsburgh School of Nursing, and the University of Pittsburgh School of Nursing Maternal/Child Health Hub.

### Competing interests

The funders had no role in the design of the study or decision to publish. As such, the authors declare no conflict of interest.

### Authors contributions

Lacey Heinsberg: Conceptualization, Funding acquisition, Methodology, Project administration, Resources, Supervision, Writing – original draft, Writing – review & editing; Miracle Loia: Conceptualization, Methodology, Project administration, Writing – original draft, Writing – review & editing; Susie Tasele: Methodology, Writing – review & editing; Kima Faasalele-Savusa: Methodology, Project administration, Supervision, Writing – review & editing; Jenna Carlson: Funding acquisition, Methodology, Writing – review & editing; Scott Anesi: Resources, Supervision, Writing – review & editing; Katie Desobry: Project administration, Writing – review & editing; Efren Yuchongco: Resources, Writing – review & editing; Brian Guevara: Resources, Writing – review & editing; Aigaeiva Sesaga: Resources, Writing – review & editing; Alice Iloilo: Resources, Writing – review & editing; Va’atausili Tofaeono: Resources, Writing – review & editing; Kaylynn Bryan: Methodology, Project administration, Writing – review & editing; Tina Tauasosi-Posiulai: Methodology, Writing – review & editing; Erin E. Kershaw: Funding acquisition, Methodology, Writing – review & editing; Yvette Conley: Conceptualization, Funding acquisition, Writing – review & editing; Daniel Weeks: Conceptualization, Funding acquisition, Methodology, Supervision, Writing – review & editing; Nicola L. Hawley: Conceptualization, Funding acquisition, Methodology, Project administration, Resources, Supervision, Writing – review & editing; Bethel Muasau-Howard: Conceptualization, Methodology, Project administration, Supervision, Writing – review & editing.

### Acknowledgements

We thank the participants—both current and future—for their time and contributions to this work. We are deeply grateful to the Lyndon B. Johnson (LBJ) Tropical Medical Center Research Oversight Committee and hospital staff for their invaluable feedback on the development of this protocol and logistical considerations. Special thanks to the clinical staff at LBJ who support the study and assist with cord blood collection—your collaboration is essential to the success of this work. We also extend our heartfelt appreciation to the prenatal care clinics at LBJ and the Department of Health, and to their staff, for being so welcoming, kind, and supportive. Your generosity and partnership make this study possible. We would also like to thank Sydney Harris, Nicole Bender, and Michelle Heller from the University of Pittsburgh School of Nursing, Department of Health Promotion and Development, for their incredible support throughout the study—including help with supply procurement, payment setup, and other essential infrastructure. Finally, we are grateful to the American Samoa Community Cancer Coalition (ASCCC) for their partnership and support in expanding and sustaining the local infrastructure that enables this work. ASCCC is a non-profit organization that has operated for the last 20 years in developing innovative ways to address the cancer burden. This has included obtaining a U24 to create the Indigenous Samoan Partnership to Initiate Research Excellence (INSPIRE) to build research capacity and assess functional health literacy (72). This led to the first National Institutes of Health R01 study awarded in American Samoa titled *Puipui Malu Manatu* – protecting memories – a study to determine Alzheimer’s Disease and Related Dementia prevalence.

### Declaration of generative AI and AI-assisted technologies

In the creation of this work, ChatGPT 4o was utilized to provide feedback on this paper, aiming to enhance readability, language, and flow. After using this tool, the authors carefully reviewed and edited the output and take full responsibility for the content of the publication.

## Notes

### Competing Interest Statement

The authors have declared no competing interest.

### Funding Statement

Yes

### Author Declarations

This study has received Institutional Review Board (IRB) approval from both the University of Pittsburgh and Yale University, with the University of Pittsburgh serving as the IRB of record through a multi-site IRB agreement (STUDY24020055). Local and territorial approval has been granted by the American Samoa IRB. In addition, the study has been approved by the Lyndon B Johnson Tropical Medical Center (LBJ) Research Oversight Committee for all hospital-related activities.

